# One-Step Rapid Quantification of Serum Neutralizing Antibody after COVID-19 Vaccination by a High-Throughput Nanoplasmonic Sensor Platform

**DOI:** 10.1101/2021.04.21.21255838

**Authors:** Liping Huang, Ying Li, Luo Changyou, Nadia Touil, Hicham el Annaz, Youqian Cheng, Shaoqi Zeng, Tang Dang, Jiawei Liang, Wenjun Hu, Hao Xu, Jiasheng Tu, Yan Shen, Gang L. Liu

## Abstract

The COVID-19 vaccination efficacy depends on serum production level of the neutralizing IgG antibody (NA) specific to the receptor binding domain of SARS-Cov-2 spike protein. Therefore, a high-throughput rapid assay to measure the total SARS-CoV-2 NA level is urgently needed for COVID-19 serodiagnosis, convalescent plasma therapy, vaccine development, and assessment. Here, we developed a nanoplasmonic immunosorbent assay (NanoPISA) platform for one-step rapid quantification of SARS-CoV-2 NAs in clinical serum samples for high-throughput evaluation of COVID-19 vaccine effectiveness. The NanoPISA platform enhanced by the use of nanoporous hollow gold nanoparticle coupling was able to detect SARS-CoV-2 NAs with a limit of detection of 0.1 ng/mL within 15 min. The one-step NanoPISA for SARS-CoV-2 NA detection in clinical specimens yielded good results, comparable to those obtained in the gold standard seroneutralization test and the surrogate virus neutralizing ELISA. Collectively, our findings indicate that the one-step NanoPISA may offer a rapid and high-throughput NA quantification platform for evaluating the effectiveness of COVID-19 vaccines.

---

The current COVID-19 pandemic, triggered by novel severe acute respiratory syndrome coronavirus 2 (SARS-CoV-2), has started in late 2019.^1-2^ The rapid spread and exponential growth of COVID-19 infections are causing health and economic havoc on a global scale, exceeding the limits of the existing healthcare and intensive care unit capacities.^3-4^ Many parts of the world have been under lockdown to curb the viral transmission.^5^ Most epidemiologists believe that SARS-CoV-2 virus will continue to spread for a long time.^6-7^ The duration of the spread of the COVID-19 epidemic mainly depends on the likelihood and duration of the immunity to the SARS-CoV-2 virus.^8^ Under such circumstances, the development of an effective vaccine and therapeutics is widely recognized as the most economical and effective way to control and ultimately end the SARS-CoV-2 virus epidemic. Thus, quantitative means to efficiently evaluate COVID-19 vaccine effectiveness are greatly needed.

Spike (S) proteins are the major surface antigens expressed on the SARS-CoV-2 virus surface.^9-10^ The receptor binding domain of the SARS-CoV-2 spike protein (S-RBD) can specifically attach to the host receptor angiotensin-converting enzyme 2 for viral entry into cells.^11-12^ Thus, S-RBD is one of the most preferred targets for making vaccines or therapeutics against SARS-CoV-2.^13-14^ The SARS-CoV-2 neutralizing antibodies (NAs) produced in response to most vaccines would effectively target the S-RBD of the SARS-CoV-2 virus and block viral entry.^15^ Currently, many SARS-CoV-2 vaccines are either in emergency use or undergoing phase II or phase III clinical trials worldwide.^16^ The traditional and gold standard method for evaluating vaccine effectiveness is the seroneutralization test, which is accurate but time-consuming and labor-intensive.^16^ It usually takes 2 to 4 days to complete the evaluation, which greatly impedes the evaluation of the large-scale population vaccination effect.^17^ Therefore, there is a pressing need for a simple, rapid and high-throughput alternative method to evaluate the level of antibodies produced after vaccination.^17-18^

Currently, active efforts are being undertaken in both the biomedical industry and basic academic research to find new diagnostic solutions and develop point-of-care devices to detect SARS-CoV-2 specific antibodies. For instance, the enzyme-linked immunosorbent assays (ELISAs),^19-20^ chemiluminescence assays,^21-22^ and lateral flow immunoassays (LFIAs)^23^ have become recently available for the detection of SARS-CoV-2 antibodies. Although ELISA and chemiluminescence assays are accurate and sensitive, the time-consuming and complex multi-step operations complicate application of these tests in a cost-effective and labor-saving way. Moreover, LFIA shows great superiority in point-of-care detection of SARS-CoV-2 antibodies because of its simple operation and rapid detection speed. In contrast, the conventional LFIA uses colloidal gold nanoparticles (GNPs) as the label probe and suffers from low throughput and sensitivity. In recent years, a novel detecting technology based on the nanoplasmonic biosensors has been developed that was combined with the scalability of detection schemes, low-cost operation, and capability for high throughput measurements, which made it very promising in point-of-care rapid detection applications.^24-26^ In addition, GNPs, due to their excellent surface plasmon resonance (SPR) properties, were widely used to enhance the detection sensitivity of the nanoplasmonic biosensor.^27-28^ Previously, we have also reported a low-cost GNP-enhanced-nanoplasmonic sensor that allowed for one-step rapid detection and quantification of the SARS-CoV-2 pseudovirus.^29^ Nanoporous hollow gold nanoparticles (NHGNPs) are core-shelled gold nanostructures with a hollow interior.^30-31^ Compared to other nanoparticles with different morphologies, NHGNPs exhibit advantages of stronger SPR properties, larger surface area, lower density, higher stability in aqueous solution, and higher sensitivity, which are extremely conducive for biomedical applications.^32-34^ Therefore, we hypothesized that a combination of nanoplasmonic nanocup sensor and NHGNPs might deliver a stronger SPR effect, which, in turn, would enable higher detection sensitivity.

To validate this hypothesis, we developed a high-throughput nanoplasmonic sensor platform for one-step rapid quantification of COVID-19 antibodies to evaluate the large-scale vaccine effectiveness (Schematic 1). The low-cost nanoplasmonic sensor platform uses an antigen sandwich method, in which the antigen S-RBD protein is used to detect total SARS-CoV-2 NAs produced by the human body. The NanoPISA can achieve high throughput detection without signal amplification and washing procedures within 15 min. The NHGNP-coupled NanoPISA platform was able to detect SARS-CoV-2 NAs in serum with an enhanced sensitivity and the limit of detection (LOD) of 0.1 ng/mL. In addition to their superior stability in serum, the NHGNPs have much larger surface area and stronger plasmon resonance than conventional colloidal GNPs, so they support higher protein density and larger resonance coupling when binding to antibodies and then to the nanoplasmonic device surface. The detection results of our developed sensor strongly correlated with those achieved by the standard ELISA (R^2^ = 0.995). Moreover, the performance of the sensor in the clinical setting was further validated by testing clinical serum specimens from 139 people that received vaccination in two nations-China and Morocco. The NanoPISA results were compared with the result of both the seroneutralization (SN) test and the surrogate virus neutralizing ELISA results. The NanoPISA exhibited over 96% sensitivity and 90% specificity. Therefore, we believe that the ultrasensitive SARS-CoV-2 detection of NAs by using NanoPISA will be very useful for the evaluation of the large-scale vaccine effectiveness and prevention of COVID-19 and other mass infectious diseases.

**Scheme 1.**
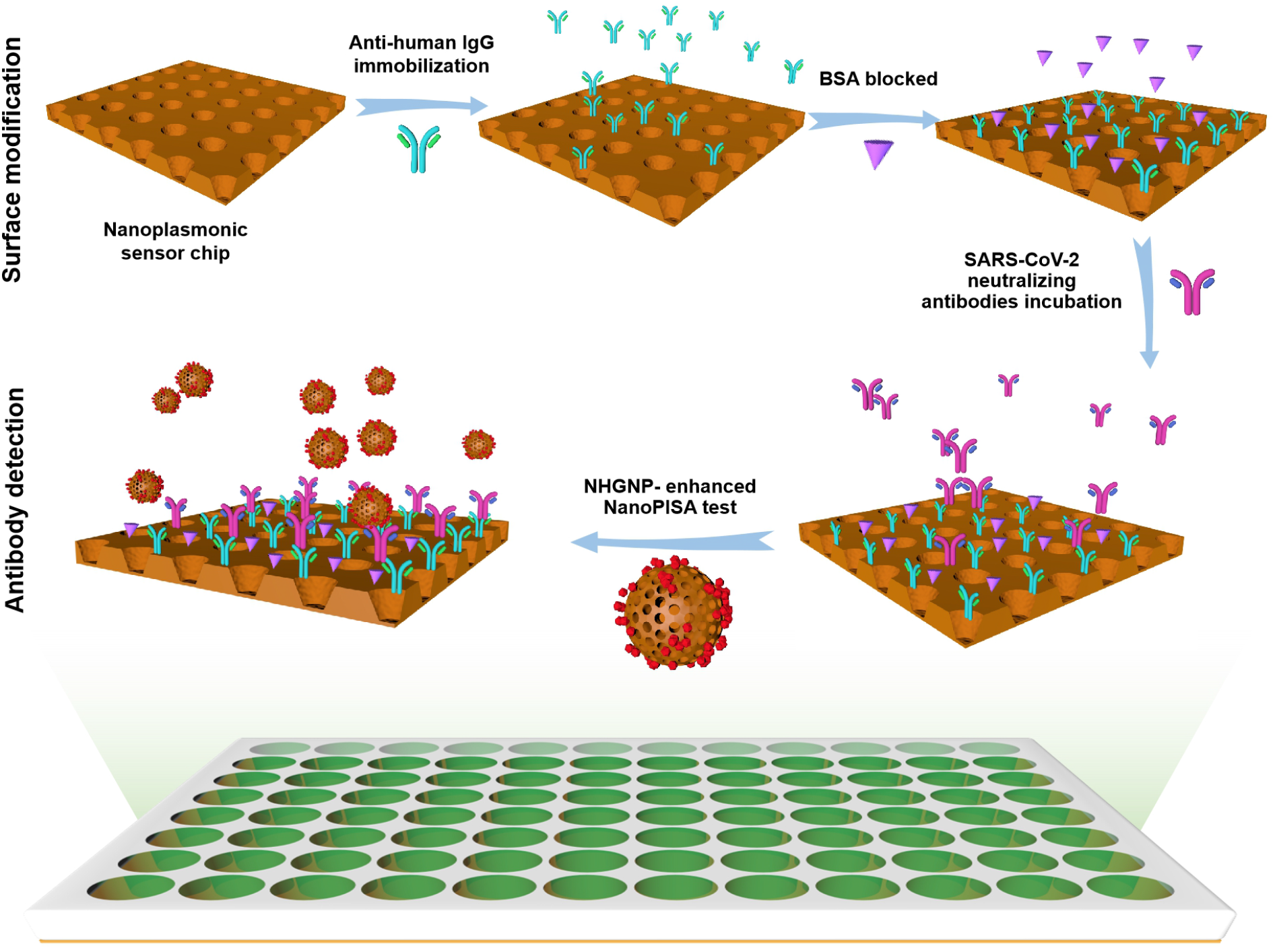
One-step rapid quantification of the total SARS-CoV-2 NAs with the NHGNP-enhanced coupled NanoPISA platform.

## RESULTS AND DISCUSSION

### Characterization of the nanoplasmonic sensor chip and NHGNPs

In this study, the periodic nanocup array sensor chip was fabricated on a polymer substrate with a diameter of 200 nm, depth of 500 nm, and periodicity of 400 nm, in which titanium (Ti) and silver (Ag) layers were 9 nm and 70 nm in thickness, respectively. It can be clearly observed that the nanocup array exhibited high uniformity in the scanning electron microscope image (Figure 1A). The sensor chip surface had distinct colors in the media with different refractive indices (RIs), such as air (RI = 1.0) and water (RI = 1.3), which shows that the sensor chip has superior detection sensitivity (Figure 1A).

**Figure 1.**
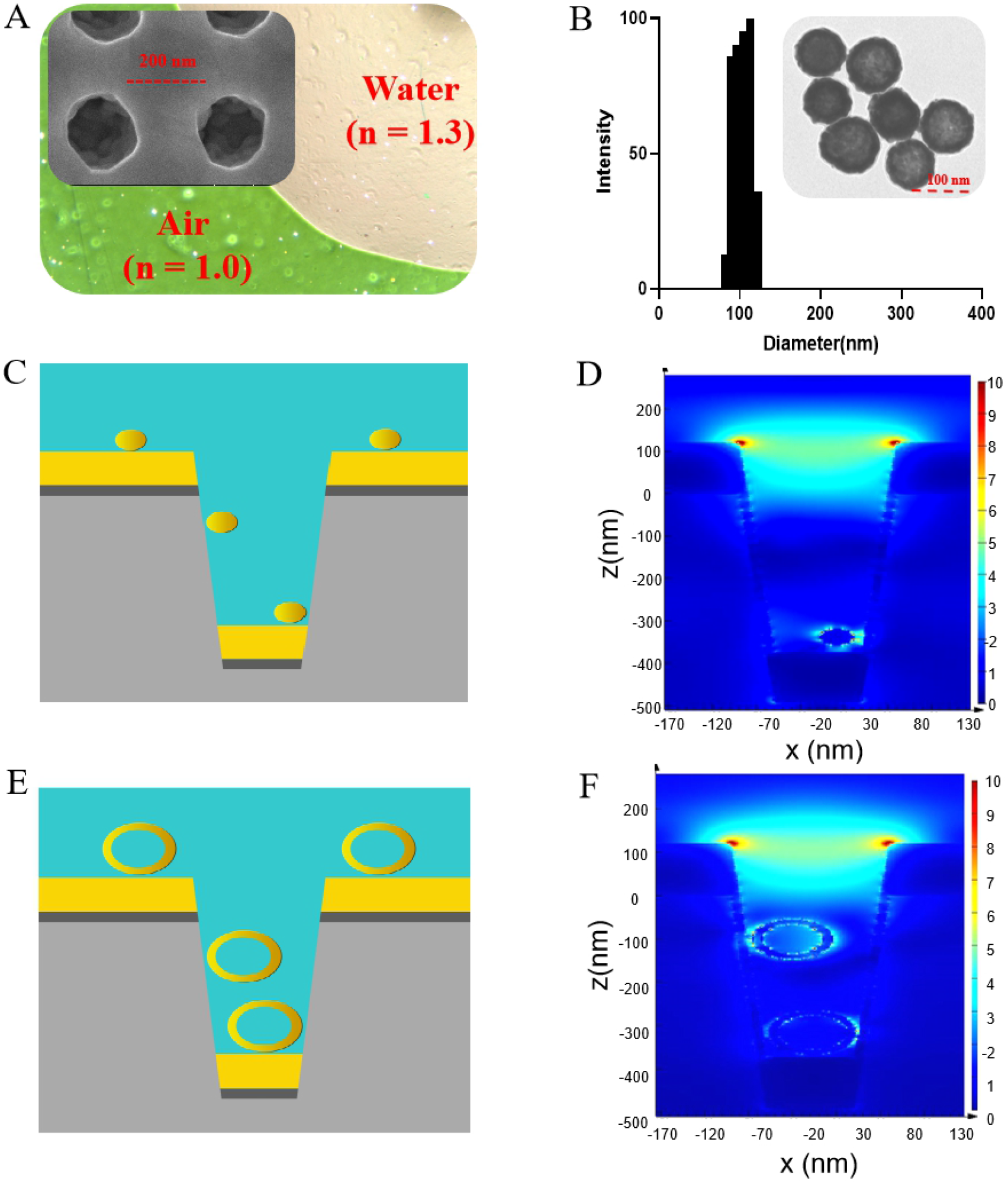
Characterization and FDTD simulation of the NHGNP enhancement mechanism on the nanoplasmonic sensor chip. (A) Transmission microscopy image shows that air and water on the device surface exhibit different colors, green and far red pink, respectively. Scanning electron microscopy image shows the replicated nanocup array (insert). (B) Size distribution and the TEM image (insert) of the NHGNPs. (C) Simulation model of colloidal GNPs coupled to the nanocup plasmonic sensor. (D) 3D-FDTD simulated electric field distributions of the colloidal GNPs coupled to the nanocup sensor. (E) Simulation model of NHGNPs coupled to the nanocup plasmonic sensor. (F) 3D-FDTD simulated electric field distributions of NHGNPs coupled to the nanocup sensor.

The NHGNP particle size was 107.2 nm with the polydispersity Index of 0.163 (Figure 1B). The synthesized NHGNPs had a negative charge (−42.48 mV) on the surface, which maintained quite a stable state because of the electrostatic repulsion caused by the adsorption of citrate ions (Figure S1). Transmission electron microscopy (TEM) images revealed that the morphology of the typical NHGNP was polyporous and hollow (Figure 1B). Moreover, the solution of NHGNPs had an effective absorption peak around 620 nm to match better the resonance peak of the nanoplasmonic sensor chip (Figure S2).

### Finite-difference time-domain (FDTD) simulations

Many studies have reported that a significant improvement in SPR performance could be achieved by coupling GNPs to the nanoplasmonic nanocup arrays, which was attempted for the generation of denser hot spots when the plasmonic GNPs were assembled on the surface of nanocups.^27, 35-36^ GNPs in the range of 20– 40 nm in diameter were generally recommended for use in biological detection procedures due to their sufficient stability and immune reactivity.^37-39^ However, only large GNPs that fit in the nanocup size can effectively help to increase the intensity of the localized electric field and improve the detection sensitivity.^40-41^ Thus, NHGNPs with higher stability and larger diameter (∼100 nm) will be a potential strategy to improve the SPR effect on the nanocup sensor chip. A quick verification of the above idea was demonstrated using the FDTD method. As expected, a stronger electric field and absorption spectra was confined in the inner cavity of the nanocup, when the NHGNPs were assembled on the nanocup array compared to colloidal GNPs (Figure 1F–H, Figure S3).

### Theory of SARS-CoV-2 NA detection by NanoPISA

The technique of the one-step rapid quantitative detection of SARS-CoV-2 NAs was based on a GNP-enhanced coupled resonance nanoplasmonic sensor with the extraordinary optical transmission effect. The coupling effect between GNPs and the nanocup array chip can significantly enhance the detection sensitivity. NHGNPs have peculiar plasmonic properties and a large exposed surface area, which attracted great scientific interest in the field of chemical and biological detection sensors.^42^ The maximization of the sensor sensitivity is achieved by the extraordinarily high structural stability of ultrafine NHGNPs. In this system, we utilized GNP-based nanoplasmonic sensor chips to quantify NAs to SARS-CoV-2. The highly specific S-RBD, which is a key target of host NAs and an important factor in vaccine design, can be used to detect total NAs in human serum. The detection schematics is illustrated in **Figure 2**, the mouse anti-human IgG solution was coated on a low-cost ultrasensitive biosensor integrated into the standard 96-well plate. When serum positive for SARS-CoV-2 NAs and the GNP-labeled S-RBD protein reagent are added successively, the target SARS-CoV-2 NAs in the serum will initially specifically bind to the GNP-labeled S-RBD and then will be captured by the anti-human IgG on the 96-well chip surface, forming the sandwich immune conjugate particles of protein-antibody-antibody. As a result, the immune conjugate particles will generate SPR effect via the ultrasensitive plasmonic biosensor chip and change optical density (OD) at a specific wavelength in the microplate reader. The relative OD value is proportional to the SARS-CoV-2 NAs concentration in serum. In addition, the NHGNP-enhanced coupled resonance nanoplasmonic sensor detection platform enables to improve the sensitivity and shorten the detection time to 15 min.

**Figure 2.**
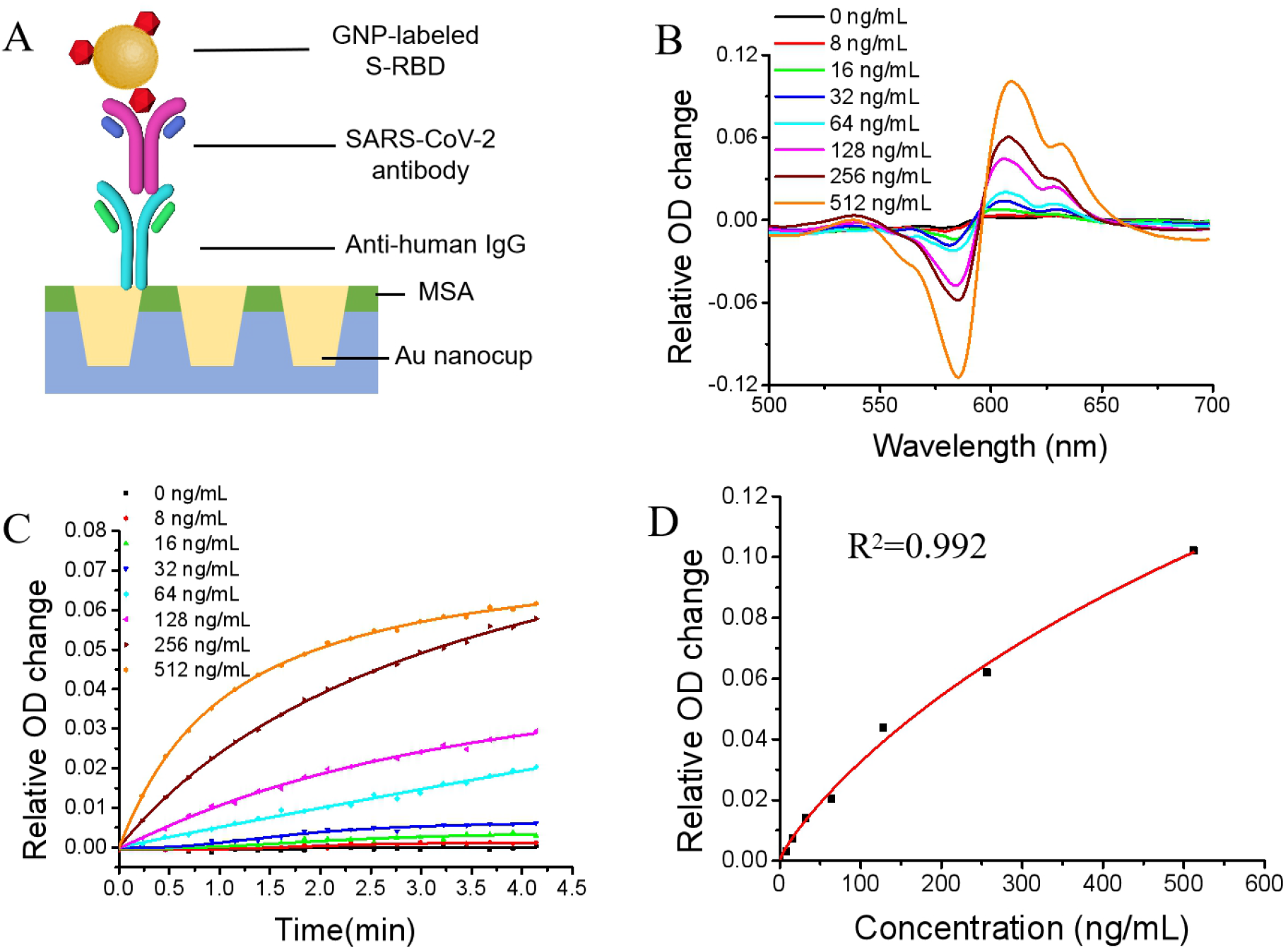
Label-free detection of the total SARS-CoV-2 NAs with colloidal GNP-enhanced NanoPISA. (A) Schematic diagram of the NanoPISA for the determination of SARS-CoV-2 Nas with colloidal GNP enhancement. (B) The differential spectra of SARS-CoV-2 NAs at a concentration range from 8 to 512 ng/mL at 590 nm and 610 nm resonant wavelengths. (C) Dynamic binding curves of SARS-CoV-2 NAs at a concentration range from 8 to 512 ng/mL at 610 nm resonant wavelength. (D) Standard curve for SARS-CoV-2 NAs (R^2^ = 0.992).

### Colloidal GNP-enhanced NanoPISA for quantitative determination of SARS-CoV-2 NAs

The developed GNP-enhanced NanoPISA biosensor was able quantitatively detect SARS-CoV-2 NAs based on the GNP-labeled S-RBD protein (Figure 2A). The NanoPISA was effective for rapid and high-throughput (96-wells) antibody testing due to its much shorter detection time (within 15 minutes in total) compared to that of ELISA detection (lasts for hours). To validate the feasibility of the GNP-enhanced NanoPISA biosensor, 50 μL of solutions containing different concentrations of SARS-CoV-2 NAs (from 0 to 512 ng/mL in 10 mM phosphate-buffered saline [PBS] with 1% bovine serum albumin [BSA]) and 10 μL of the solution with GNP-labeled S-RBD protein were successively added into each well of the anti-human IgG modified sensor chip integrated with a microwell plate for the detection via a generic microplate reader. The diluted solution without SARS-CoV-2 NAs was used as the control blank group. As shown in Figure 2B, the absorption spectra of solutions with various concentrations of SARS-CoV-2 NAs were quite different. The specific resonant wavelength of the colloidal GNP-coupled nanocup arrary chip could be seen at 590 nm and 610 nm, and the opposite variations were observed at 590 nm and 610 nm with respect to the different concentrations of SARS-CoV-2 NAs. The real-time dynamic binding curves of the sandwich immune conjugate particles interaction at resonant wavelength 610 nm are shown in Figure 2C. The relative OD intensity positively correlated with SARS-CoV-2 NAs concentration, whereas the dynamic curve of the control blank group showed almost no change. Furthermore, the difference between the relative OD change at 610 nm and 590 nm was obtained from the differential absorption spectra at 15 min. The standard curve of the relative OD value in the colloidal GNP-enhanced NanoPISA relative to the concentration of SARS-CoV-2 NAs in the range from 0 to 640 ng/mL is shown in Figure 2C. The standard curve is a sigmoid curve obtained using a four-parameter logistic model, and the correlation coefficient (R^2^) was 0.992. The LOD of the colloidal GNP-enhanced NanoPISA for the SARS-CoV-2 NA detection was estimated to be 5 ng/mL.

### NHGNP-enhanced NanoPISA for quantitative determination of SARS-CoV-2 NAs

The excellent colloidal stability of GNPs is very important for their various applications in different fields of sciences. Unfortunately, the stability of colloidal GNPs can be easily affected by many external factors, such as solution environment and protein concentration.^43-44^ However, NHGNPs with stronger SPR properties, larger surface area, and higher stability in aqueous solutions may have higher detection sensitivity than colloidal GNPs. Based on the nanoplasmonic sensor chip plate, we developed an enhanced detection reagent with high stability of NHGNPs that enabled over 50-fold increase in the detection sensitivity (Figure 3A). Similarly, the anti-human IgG-modified sensor surface and nanoporous hollow AuNP-labeled S-RBD protein were used for rapid detection of SARS-CoV-2 NAs by one-step method as described above. In the typical differential absorption spectra of the NHGNP-enhanced sensor chip, the specific resonant wavelengths appeared at 590 nm and 650 nm (Figure 3B). The opposite variation values, proportional to the different concentrations of SARS-CoV-2 NAs, were also observed at 590 nm and 650 nm at 15 min. In addition, the dynamic curves of SARS-CoV-2 NAs at different concentrations and the blank control sample showed obvious distinction within 5 min (Figure 3C). As expected, the one-step NHGNP-enhanced NanoPISA significantly improved the detection sensitivity of SARS-CoV-2 NAs at a concentration of 0.1 ng/mL, which was 50 times more sensitive than colloidal GNP-enhanced NanoPISA. Furthermore, the standard curve of the difference between the relative OD values at 650 nm and 590 nm with respect to different concentrations of SARS-CoV-2 NAs was obtained from the differential spectra, and the 4-parameter logistic curve fitting. The R^2^ was found to be 0.998 over the range of SARS-CoV-2 NA concentrations from 0.5 to 125 ng/mL (Figure 3D).

**Figure 3.**
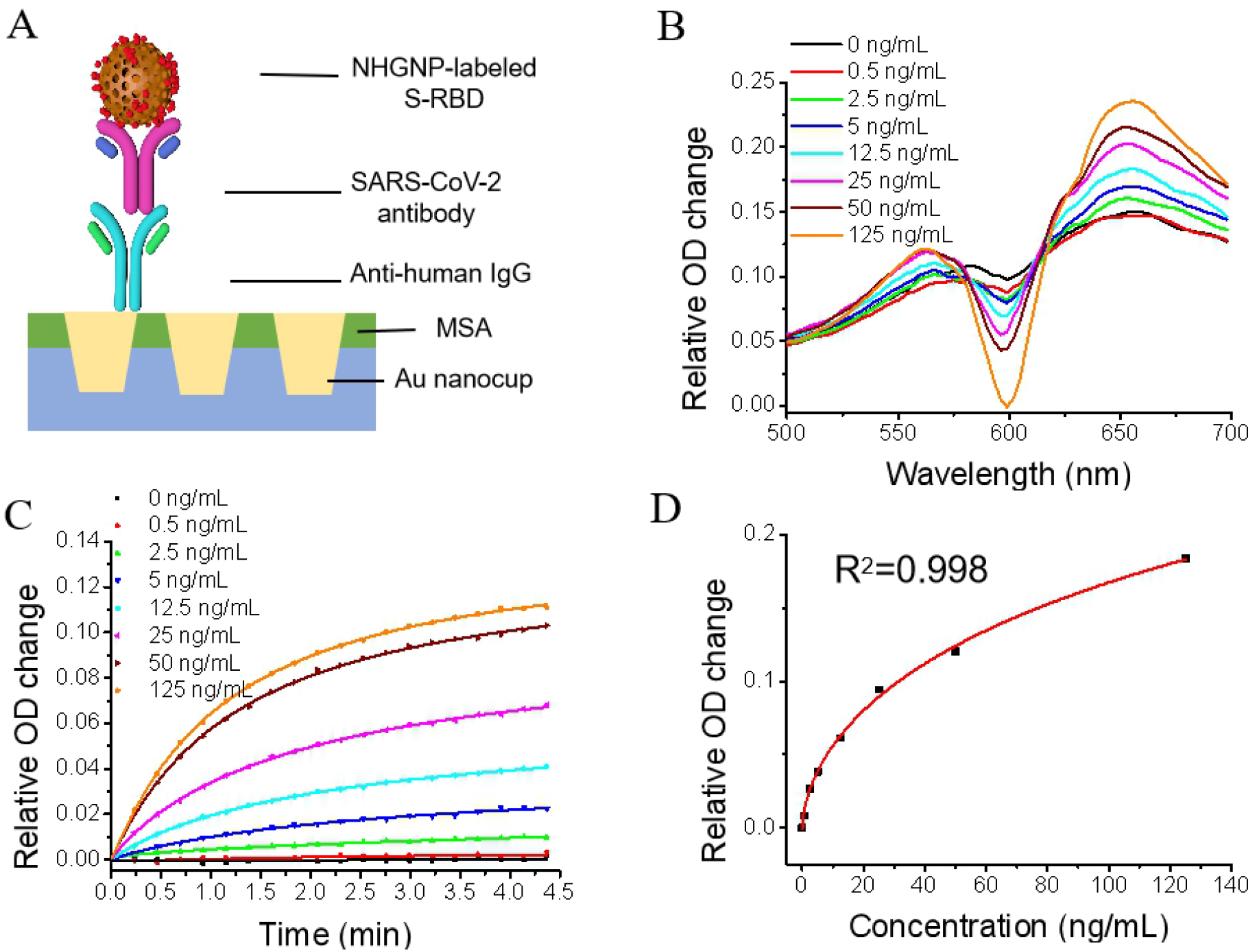
Label-free detection of the total SARS-CoV-2 NAs with NHGNP enhancement based on the NanoPISA platform. (A) Schematic diagram of the nanoplasmonic resonance sensor for the determination of SARS-CoV-2 NAs with NHGNP enhancement. (B) The differential spectra of SARS-CoV-2 NAs at different concentrations (0–125 ng/mL) at 600 nm and 650 nm resonant wavelengths. (C) Dynamic binding curves of SARS-CoV-2 NAs at different concentrations over the range from 0 to 125 ng/mL. (D) Standard curve of SARS-CoV-2 NA detection by NHGNP-labeled S-RBD (R2 = 0.998, LOD = 0.1 ng/mL).

At present, a safe and effective COVID-19 vaccine is the best way to control and ultimately end the pandemic, so the development of COVID-19 vaccines is moving at unprecedented speeds. However, the question of how to evaluate the effectiveness of the vaccine is very challenging. In this study, the proposed one-step NanoPISA platform not only detected SARS-CoV-2 NAs with ultrahigh sensitivity and effectiveness, but also significantly relieved the workload of medical personnel who analyzed numerous serum samples from vaccinated people, consequently improving the efficiency of the evaluation of vaccine effectiveness.

### Comparative analysis of the NanoPISA and ELISA for SARS-CoV-2 antibody

A schematic comparison of the one-step NanoPISA and multistep ELISA with estimated time spent on each step for single sample detection is shown in Figure S4. SARS-CoV-2 antibodies were diluted in a buffer to obtain a concentration range of 10–640 ng/mL (the same concentration range applied for ELISA). The resulting sensor calibration curve is displayed in Figure S5. Each data point was averaged from two independent measurements. Figure S6 shows the corresponding SARS-CoV-2 antibody standard curve measured by ELISA. A very good correlation between NanoPISA and ELISA was achieved for the detection of SARS-CoV-2 antibody dissolved in the buffer and the fitting R^2^ was 0.995 (Figure S7). To determine the accuracy of the proposed AuNP-enhanced NanoPISA for SARS-CoV-2 antibody detection, we analyzed three samples of SARS-CoV-2 antibody in buffer with different concentrations from 20 ng/mL to 160 ng/mL using NanoPISA and ELISA. The average value and accuracy rate for each sample were calculated. As shown in Table S1, the accuracy rate in the same sample is between 80% and 120%, indicating that both the NanoPISA and ELISA have good stability and accuracy. The NanoPISA was optimized to use relatively small sample volumes (∼50 □ L) while maintaining high sensitivity in comparison with ELISA, which can be attributed to the use AuNP-enhanced coupled resonance nanoplasmonic sensor chip. Another major advantage of the NanoPISA technique is that SARS-CoV-2 antibody can be quickly detected with higher throughput compared with ELISA. The high throughput and detection speed of the NanoPISA allowed for all sample tests to be finished within 15 min, whereas it took over 3 h to complete the conventional ELISA.

### Validation of NanoPISA using real clinical samples

In order to confirm the diagnostic accuracy of our developed NanoPISA technique, a scale-up clinical serum sample test was performed at the Hôpital Militaire d’Instruction Med V Rabat. Serum samples were obtained from 139 volunteers without SARS-CoV-2 infection before and after Covid-19 vaccination. The detection of SARS-CoV-2 NA level in each clinical serum sample was performed as described above, using only 50 µL of diluted samples for each measurement. The measurements were done in a blind manner, where the true NAs concentration level is not known. After the completion of the clinical tests, the results collected from the NanoPISA were compared with those of the previous surrogate virus neutralizing ELISA (Table 1) as well as the SN test (Table 2).

**Table 1.**
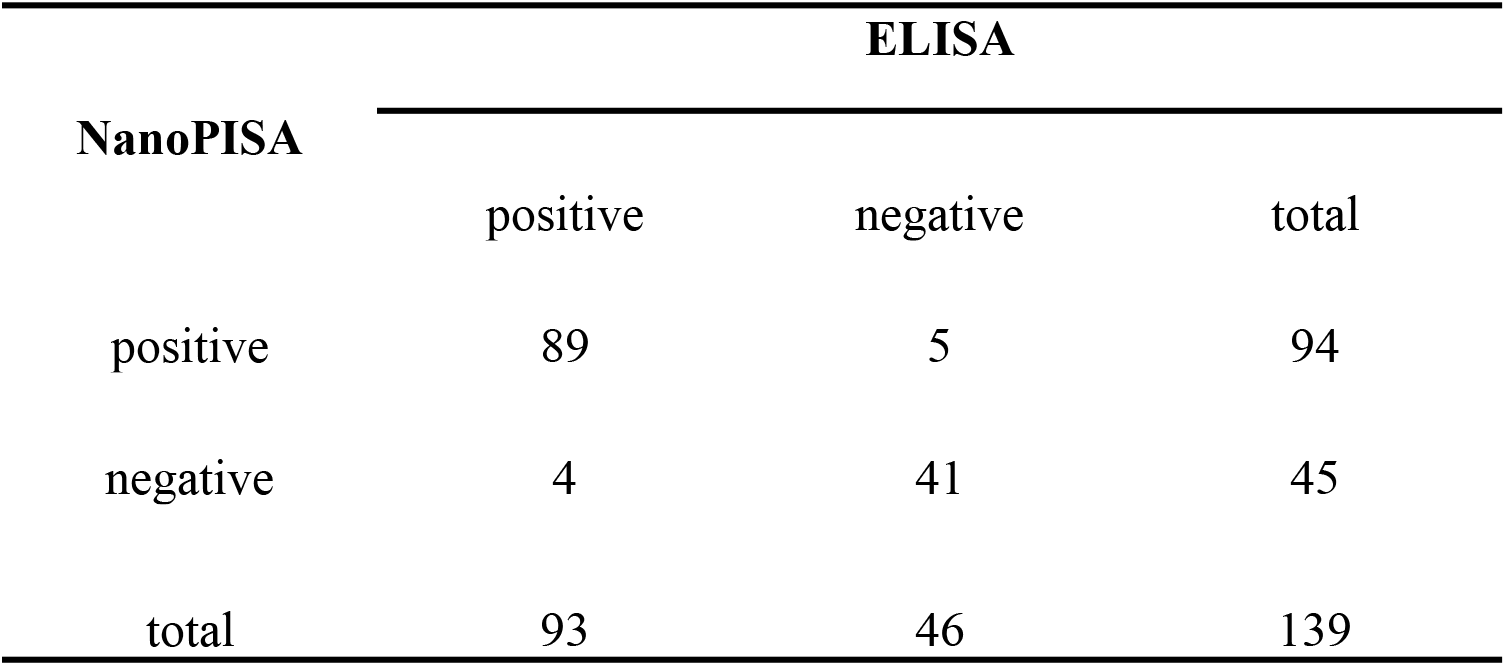
χ^2^ Test for Diagnostic Results of the Developed NanoPISA and ELISA.

**Table 2.** χ^2^ Test for Diagnostic Results of the Developed NanoPISA and SN Test.

| NanoPISA | SN test |  |  |
| --- | --- | --- | --- |
|  | positive | negative | total |
| positive | 34 | 6 | 40 |
| negative | 3 | 25 | 28 |
| total | 37 | 31 | 68 |

NanoPISA results of the 139 clinical serum samples were first compared with the results of the surrogate virus neutralizing ELISA. The percentage of NanoPISA NA-positive samples was similar to that seen with ELISA. NanoPISA exhibited 95.7% sensitivity and 89.1% specificity compared to those of ELISA (Table 1). In addition, the χ^2^ test was used to compare the results between NanoPISA and ELISA, which showed that the *P*-value of the McNemar test was 1.0, and the value obtained with the kappa test was 0.898 (*P* < 0.001). The results of the NanoPISA were not significantly different from those of ELISA. In addition, SARS-CoV-2 NA levels in 68 clinical serum samples also showed very strong concordance between the NanoPISA and SN test results, with the kappa coefficient of 0.604 (*P* < 0.001). The sensitivity and specificity of the NanoPISA compared to those of the SN test were 91.9% and 80.7%, respectively. Thus, there is very high concordance between the results of NanoPISA and of two alternative immunoassays, suggesting robust NanoPISA performance.

## CONCLUSION

In this study, we have described a rapid and high throughput nanoplasmonic sensor-based assay, or NanoPISA, for SARS-CoV-2 NAs. A 96-well biosensor chip plate device was pre-coated with mouse anti-human IgG and with NHGNP-labelled S-RBD, the samples were subsequently added for directly measuring SARS-CoV-2 NA levels in blood serum. The unique nanoplasmonic mechanism through NHGNPs coupled to the nanocup sensors enable highly sensitive and rapid SARS-CoV-2 NAs detection in one step. When diagnosing the clinical serum samples, there was very strong concordance between the results of the NanoPISA for SARS-CoV-2 NAs and those of the conventional ELISA and SN assay. In addition, compared with the conventional ELISA and SN assay, the NanoPISA shortened the total detection time from days and hours to within only 15 min and improved the testing capacity to simultaneous high throughput detection of up to 96 samples. We therefore conclude that the NanoPISA platform-based low-cost high-throughput devices could rapidly evaluate the large-scale vaccine effectiveness, promoting vaccine development and ultimately ending the spread of SARS-CoV-2 virus epidemic.

## MATERIALS AND METHODS

### Materials

Hexylsilane, 6-mercapto-1-hexanol (MCH), mercaptosuccinic acid (MSA), 1-ethyl-3-(dimethylaminopropyl) carbodiimide (EDC), N-hydroxysuccinimide (NHS), bovine serum albumin (BSA), ethanolamine and phosphate-buffered saline (PBS) buffer were purchased from Sigma-Aldrich. Trisodium citrate dehydrate and NaBH_4_ were purchased from Nanjing Chemical Reagent Co.Ltd.. CoCl_2_ were purchased from Guangdong Guanghua Sci-Tech Co.Ltd.. HAuCl_4_ were purchased from Sinopharm Chemical Reagent Co.,Ltd.. SARS-CoV-2 Spike Protein RBD, mouse anti-human IgG (catalog no. V90401) was purchased from Nanjing Genscript Co., Ltd.. SARS-COV-2 RBD-mAbs (GHMA 105-1, GHMA 105-2) was purchased from Goodhere Biological Technology Co., Ltd.. All chemicals were used as obtained without any further purification.

Serum samples were obtained from the Hôpital Militaire d’Instruction Med V Rabat and stored at −80 °C until use. This study was approved by the Ethics Committee of the Science and Technology Department of Huazhong University of Science and Technology (Certificate #: S1029).

#### Fabrication and characterization of the nanocup array sensor chip

The nanoplasmonic sensor chip was fabricated by the replica molding technique with a mold. The original mold was composed of a tapered nanopillar array and the periodicity, height, and width of the nanocone were 400, 500, and 200 nm, respectively. The UV-curable polymer was evenly spread on the mold and placed on a polyethylene terephthalate sheet to produce the polymeric nanocup array structure. Then, 9 nm of titanium and 70 nm of gold were subsequently deposited on the polymeric nanocup array in an electron beam evaporator. Then the sheet was cut to 13 cm × 8.5 cm and glued to an open-bottom 96-well plate made by a 3D printer (Objet30 Prime™, Stratasys Ltd).

#### Surface functionalization

Chip surface modification was prepared as described in our previous study._19_ In brief, 100 μL of 50 mM sulfhydryl succinic acid solution was added into a homemade 96-well chip plate and kept for 1 h at room temperature. After cleaning the chip well with deionized water twice, 100 μL of 400 mM EDC and 100 mM NHS (in PBS) was added and left for 30 min to activate the carboxyl group of sulfhydryl succinic acid at room temperature. Next, the chip wells were rinsed with deionized water and 50 μL of an 8.0 μg/mL anti-human IgG PBS solution was immediately immobilized on the activated chip wells for 4 h at room temperature. Then, the chip wells were blocked with 100 μL of a 60 μg/mL BSA solution and then with 100 μL of a 10% ethanolamine solution for 30 min at room temperature, respectively. Finally, the functionalized chips were gently rinsed with deionized water twice and stored at 4 °C for further use.

#### Preparation of GNPs and GNP-labeled S-RBD protein

Colloidal GNPs were prepared by a sodium citrate reduction method.^45-46^ Briefly, 300 mL of 1 mM of HAuCl_4_ was heated until it boiled and then 15 mL of 75 mM citrate was added to the boiling solution. A dark red color solution was obtained, which was boiled continuously for another 15 min and then allowed to cool down to room temperature.

The typical procedure of labeling the S-RBD protein with colloidal GNPs was employed as described in our previous work.^29^ First, the pH of 1.5 mL of the colloidal GNP solution was adjusted to pH 7.4 by 0.1 M K_2_CO_3_ solution. Then, 4.9 □ L of 0.93 mg/mL S-RBD protein solution in PBS was added to the colloidal AuNP solution and incubated with AuNPs for 15 min. After blocking with 22.5 μL of the blocking solution containing PEG 2W (10%, w/v) for 15 min, the colloidal GNP suspension was centrifuged at 7,500 rpm for 22 min. Finally, the precipitate of the GNP-labeled S-RBD protein was resuspended in 530 μL of stabilizing buffer (20 mM Tris, pH 9.2, 0.3% sucrose, 0.05% PEG 20000) and stored at 4 °C for further experiments.

#### Preparation of NHGNPs and NHGNP-labeled S-RBD protein

Three hundred milliliters of ultrapure water, 6 mL of 0.05 M sodium citrate, and 300 μL of a 0.4 M CoCl_2_ solution was consecutively added to a 500 mL round-bottomed flask. After the solution mixture was stirred for 10 min in vacuum conditions, 3 mL of a 0.13 mM NaBH_4_ solution in water was added to the flask. The mixture continued to be stirred for 15 min until the solution became darker color. Then 1.5 mL of a 25 mM HAuCl_4_ solution was added to the flask and stirred overnight. Finally, 300 μL of a 20% PVP solution was added to the reaction mixture and stirred for 30 min. The final reaction solution was centrifuged for 15 min at 9,000 r/min in a high-speed refrigerated centrifuge. The supernatant was discarded, and the precipitate was resuspended with ultrapure water and stored at 4 °C for further experiments.

The morphology and size of the synthesized NHGNPs based on Co templates were characterized by transmission electron microscopy (TEM, HT7700, Japan) with a CCD camera operating at an accelerating voltage of 100 kV. The NHGNP particle size distribution was determined by dynamic light scattering on a Zetasizer Nano-ZS90 analyzer (Malvern Instruments, UK). The absorption spectra of NHGNPs ranging from 400 to 900 nm were obtained using a UV-vis spectrophotometer (Agilent 8453, USA) at 25 °C.

S-RBD protein labeling with NHGNPs was done as described in our previous work, with minor modifications as follows.^29^ Briefly, 9 □L of a 0.93 mg/mL S-RBD protein solution in PBS was added to a 0.2 mg/mL NHGNP solution and incubated with NHGNPs for 15 min. Then, 15 μL of the blocking solution containing PEG 2W (10%, w/v) was added into the NHGNP solution and incubated for 15 min. Then, the NHGNP suspension was centrifuged at 7,500 rpm for 20 min. Finally, the NHGNP precipitate was resuspended in 530 μL of the stabilizing buffer (20 mM Tris, pH 9.2, 0.3% sucrose, 0.05% PEG 20000) and stored at 4 °C for further experiments.

#### FDTD simulations

Three dimensional FDTD simulations were performed by commercial software (FDTD Solutions, Lumerical). The model had the same geometry as the actual sensor device with a periodicity of 400 nm, nanocup top diameter of 200 nm, nanocup height of 500 nm, respectively. The spheres and hollow spheres were used to simulate colloidal GNPs and NHGNPs. The background refractive index was set to 1.333 to simulate liquid environment. The sensor chips were illuminated with a plane wave from the top side (-z direction). Perfect matching layer was applied to the boundary conditions in the z axis and periodic was applied to the boundary conditions in the x axis and y axis.

#### GNP-enhanced NanoPISA for rapid detection of the total new coronavirus SARS-CoV-2 NAs

To demonstrate the capabilities of the GNP-enhanced NanoPISA platform, the SARS-CoV-2 NAs were quantitatively detected using NanoSPR combination sensors. In detail, 50 μL of solutions with different concentrations of SARS-CoV-2 NAs (in 10 mM PBS, 1% BSA) was added to each individual well of the anti-human IgG functionalized sensor chip plate. Then, 10 μL of the gold-labeled S-RBD protein solution was added to each well of the sensor chip plate. The typical original spectra and dynamic interaction kinetics of the SARS-CoV-2 NAs were recorded using a generic microplate reader at the specific resonant wavelength. After 15 min, the final spectra of solutions with different concentrations of NAs were recorded again by using a generic microplate reader.

Detection of SARS-CoV-2 NAs in human serum samples was the same as in the protocol described above, but for the sample preparation step, the plasma or serum samples required 1:100 dilution with sample diluent buffer (10 mM PBS, 1% BSA) to fall within the range of the assay. Plasma samples were obtained by centrifuging blood samples at 2,500×g and 4 °C for 15 min. The serum samples were stored at −80 °C.

In addition, the detection of the SARS-CoV-2 NAs by NanoPISA were validated by SARS-CoV-2 Antibody Assay ELISA kit (XG100H2, AbMax Biotechnology Co., Ltd.) and SARS-CoV-2 surrogate virus neutralizion ELISA kit (L00847-A, Nanjing GenScript Biotech Co., Ltd.). SARS-CoV-2 Antibody Assay ELISA was performed by incubating for 30 min with 100 μL of different concentrations of SARS-CoV-2 antibody in a shaking incubator (37 ° C, 200 rpm), washing 3 times with Washing Buffer (250 μ L/well), and adding 100 μ L of diluted enzyme-conjugated pAb solution to each well for 15 min with taping every 5 min. Wells were washed 3 times and 50 μL each of Substrate Solution A and Substrate Solution B was added for 10 min at dark, the reaction was stopped by adding 50 μ L of 0.2M H2SO4 into each well, and optical density (OD) of each well was recorded at 450 nm on a plate reader within 30 min.

SARS-CoV-2 surrogate virus neutralizion ELISA was performed by incubating the samples with the diluted HRP-RBD solution with a volume ratio of 1:1 in separate tubes at 37°C for 30 min. Add 100 µL each of the sample mixture to the corresponding wells and incubate at 37°C for 15 minutes. Wash the plate with 260 µL of 1×Wash Solution for four times. Add 100 µL of TMB Solution to each well and incubate the plate in dark at 20 - 25°C for 15 min. Add 50 µL of Stop Solution to each well to stop the reaction and read the absorbance in the microtiter plate reader at 450 nm immediately.

## Data Availability

All relevant data are available from the authors.

## ASSOCIATED CONTENT

### Supporting Information

The Supporting Information is available free of charge on the ACS Publications website. Figures of potential, absorption spectra of NHGNPs, the FDTD simulated absorption spectra of GNPs and NHGNPs coupled to the nanocup sensor, schematic comparison, calibration curves and the accuracy comparison of the NanoPISA and ELISA on SARS-CoV-2 NAs detection (PDF)

## Author Contributions

Liping Huang, Ying Li, and Changyou, Luo contributed equally to this work. Liping Huang: Conceptualization, Methodology, Investigation, Data curation, Visualization, Writing - original draft. Ying Li: Data acquisition, Investigation, Software. Changyou Luo: Data acquisition. Nadia Touil: Clinical data acquisition., Hicham el Annaz: Clinical data acquisition. Youqian Cheng: Data acquisition. Shaoqi Zeng: GNP-labled S-RBD preparation. Tang Danga: Software. Jiawei Liang: Software. Jiasheng Tu: Investigation, Resources. Hao Xu: Supervision, Funding acquisition. Yan Shen: Supervision, Funding acquisition, Project administration. Gang L. Liu: Supervision, Writing - review & editing, Project administration.

## Notes

The authors declare no competing financial interest.

## ACKNOWLEDGMENTS

This work was supported by a grant from the National Key R & D Plan of China (2020YFC0861900), the National Natural Science Foundation of China (91959107, 81972892) and Fundamental Research Funds for the Central Universities (2020kfyXGYJ111). The authors thank Xiaofeng Du, Hanlin Zhou, Hong Yang, and Hui Guo for their help with data acquisition. We would like to thank Editage (www.editage.cn) for English language editing.

