## Supplemental information for "One-Step Rapid Quantification of Serum Neutralizing Antibody after COVID-19 Vaccination by a High-Throughput Nanoplasmonic Sensor Platform"

**Supplementary Figure S1**

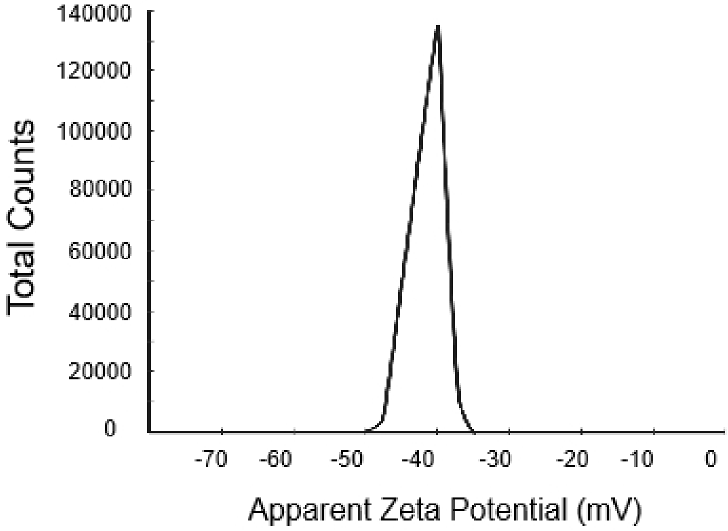


**Figure S1.** The potential of the NHGNPs.

**Supplementary Figure S2**

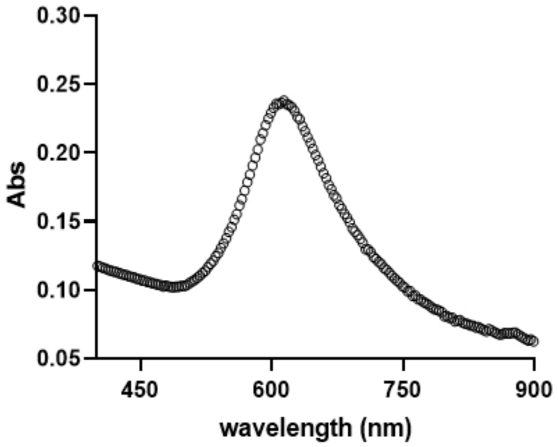


**Figure S2.** The absorption spectra of the NHGNPs.

**Supplementary Figure S3**

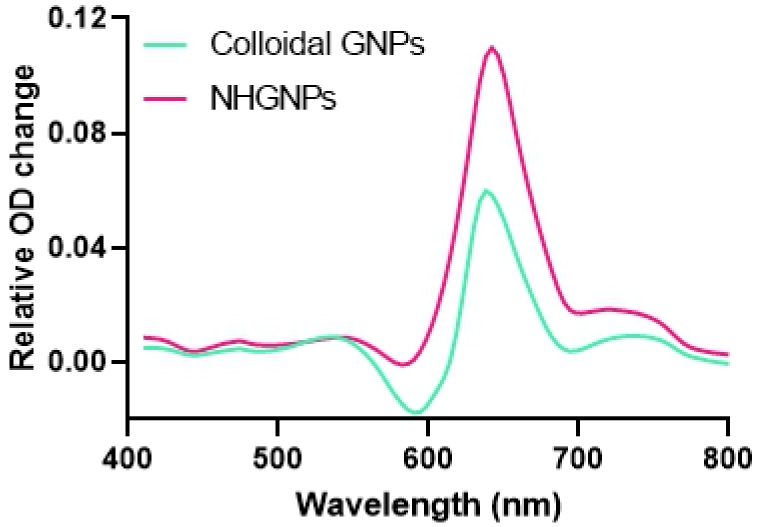


**Figure S3.** The FDTD simulated absorption spectra of GNPs and NHGNPs coupled to the nanocup sensor.

### Supplementary Figure S4

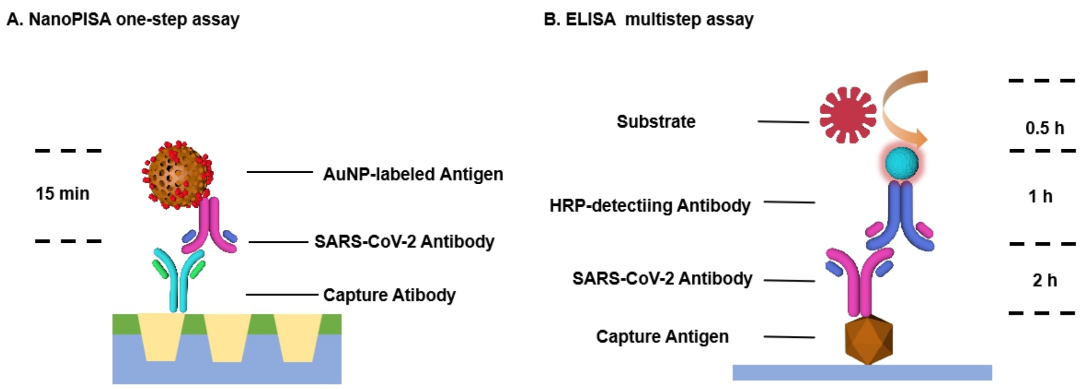


**Figure S4.** A schematic comparison of the NanoPISA one-setp assay and ELISA multistep assay with estimated time spent on each step for single-sample detection.

**Supplementary Figure S5**

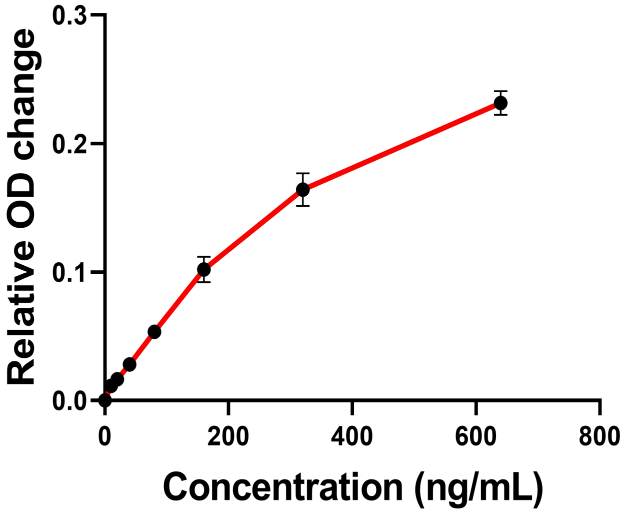


**Figure S5.** The calibration curve of SARS-CoV-2 antibody measured by NanoPISA one-setp assay.

**Supplementary Figure S6**

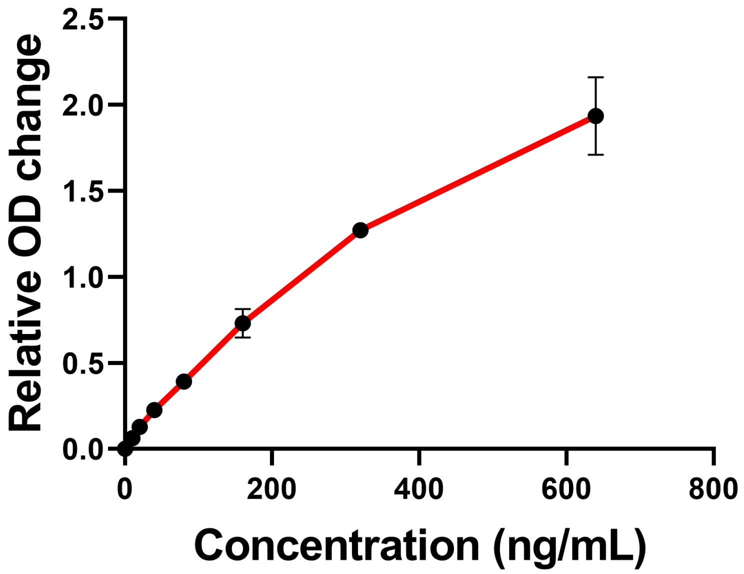


**Figure S6.** The standard curve of SARS-CoV-2 antibody measured by ELISA multistep assay.

**Supplementary Figure S7**

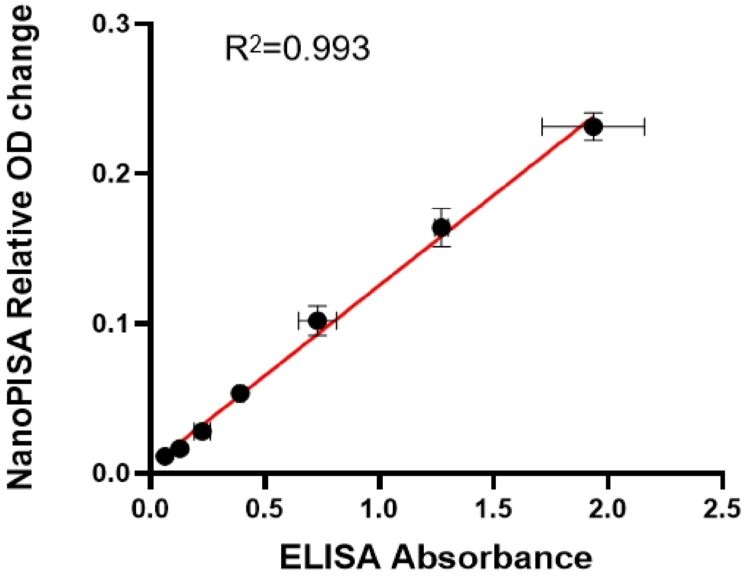


**Figure. S7.** A correlation between NanoPISA and ELISA methods in detection of SARS-CoV-2 S-RBD protein antibody with the concentrations of 10, 20, 40, 80, 160, 320, and 640 ng/mL.

**Supplementary Table S1**

**Table S1** The accuracy of the proposed AuNPs enhanced NanoPISA assay for SARS-CoV-2 antibody detection,

| | ELISA | | NanoPISA | |
|---|---|---|---|---|
| Concentration(ng/mL) | Average (ng/mL) | Accuracy(%) | Average (ng/mL) | Accuracy(%) |
| 20 | 24.09 | 120.4% | 20.19 | 100.9% |
| 80 | 82.01 | 102.5% | 74.75 | 93.4% |
| 320 | 317.90 | 99.3% | 305.98 | 95.6% |
